# Implementation of data access and use procedures in clinical data warehouses. A systematic review of literature and publicly available policies

**DOI:** 10.1101/2020.01.27.20018861

**Authors:** Elena Pavlenko, Daniel Strech, Holger Langhof

## Abstract

**Background:** The promises of improved health care and health research through data-intensive applications rely on a growing amount of health data. At the core of large-scale data integration efforts, clinical data warehouses (CDW) are also responsible of data governance, managing data access and (re)use. As the complexity of the data flow increases, greater transparency and standardization of criteria and procedures is required in order to maintain objective oversight and control. This study assessed the spectrum of data access and use criteria and procedures in clinical data warehouses governance internationally.

**Methods:** We performed a systematic review of (a) the published scientific literature on CDW and (b) publicly available information on CDW data access, e.g., data access policies. A qualitative thematic analysis was applied to all included literature and policies.

**Results:** Twenty-three scientific publications and one policy document were included in the final analysis. The qualitative analysis led to a final set of three main thematic categories: (1) requirements, including recipient requirements, reuse requirements, and formal requirements; (2) structures and processes, including review bodies and review values; and (3) access, including access limitations.

**Conclusions:** The description of data access and use governance in the scientific literature is characterized by a high level of heterogeneity and ambiguity. In practice, this might limit the effective data sharing needed to fulfil the high expectations of data-intensive approaches in medical research and health care. The lack of publicly available information on access policies conflicts with ethical requirements linked to principles of transparency and accountability.

CDW should publicly disclose by whom and under which conditions data can be accessed, and provide designated governance structures and policies to increase transparency on data access. The results of this review may contribute to the development of practice-oriented minimal standards for the governance of data access, which could also result in a stronger harmonization, efficiency, and effectiveness of CDW.

## BACKGROUND

Digitalization in health care and biomedical research has developed at a rapid pace. What previously consisted of error-prone and time-consuming manual documentation of information, often resulting in poorly structured data, has in large part been replaced by digitally supported or fully automatized processes. Despite persistent challenges, the widespread adoption of electronic health records (EHR) is one of many examples of this digital progress (1), adding to an ever-increasing amount of personal clinical data generated in routine health care delivery.

High potential is expected from the reuse of such structured clinical data for secondary research purposes, for example when applied in epidemiological and health economic research, comparative effectiveness research, or health care quality improvement (2-4). The establishment of learning health care systems (LHCS) and the development of precision medicine are further examples of promising developments (5, 6). Given an appropriate level of data quality, data-intensive research using big data analytics, machine learning, deep learning and artificial intelligence could become game changers in biomedical research and health care delivery (7).

One of the cornerstones of successful data reuse is an appropriate data infrastructure. However, due to differences in local, regional and national infrastructures, the information system landscape in the health care sector is largely characterized by heterogeneity (8). Hospitals alone, for instance, require a broad spectrum of different IT solutions, such as electronic medical records, laboratory information systems, and individual solutions for clinical research (e.g., databases, registries) at the same time. In many cases, the interoperability of these systems is limited (9). Thus, a major concern for the effective usage of clinical data even within one system is the often-fragmented data storage creating so called “data silos” (10, 11).

First developed for the industry, data warehousing has been identified as a solution to overcome this siloed infrastructure (11). Clinical data warehouses (CDW), more specifically, consolidate and integrate clinical data from various sources, such as health care data (e.g., from EHR), medical research data (e.g., from research biobanks or clinical trials) and patient-generated data (e.g., via mobile phones, smart-health applications, or wearables) (12). When fully implemented, CDW allow for a broad and real-time analysis of data at the levels of individual patients and cohorts. The vast amount of data provided by CDW is thus seen as a key resource for data-intensive approaches such as research in precision medicine and quality improvement (12-15).

However, CDW are more than a mere technical infrastructure to integrate data. CDW hold responsibility over data stewardship (16), meaning the management and oversight of data, playing a crucial role in making stored clinical data accessible and (re)usable. With the multiplying promises of data-intensive research, the governance of data access and use gains an ethical dimension whose relevance is debated internationally (4, 17, 18).

An abundance of policies, regulations and guidelines addresses the importance of sharing data and making health data accessible for research purposes, thus calling for a transparent and sustainable data access governance (19). For instance, the World Medical Association (WMA) in their Declaration of Taipei clearly demands, “Governance arrangements must include (…) criteria and procedures concerning the access to and the sharing of health data” (20).

Challenges often arise when ethical standards are implemented in practice. In the case of biobanks, for instance, a recent investigation on sample and data access governance revealed significant shortcomings: although sample and data access policies are required, their availability is limited and the criteria for access decisions outlined in the policies lack systematization and harmonization (21). This is recognized as a threat to responsible and transparent (inter-)national collaboration and thus limits the prospects of networked biobanking (22).

Regarding data governance and stewardship, biobanks and CDW share common responsibilities. So far, the scientific literature offers little information about the actual practice of structuring and handling the governance of data access in CDW. A review by Holmes et al. on data warehouse governance for distributed research networks found that details on governance were sparse. However, their review was limited to publications from the U.S. before July 2013, and the authors expected more fitting publications to emerge in the years following their publication (23).

The aim of this study was to assess the spectrum of criteria and procedures applied in data access and use governance in CDW internationally.

## METHODS

We performed a systematic review of (a) published scientific literature on CDW and (b) publicly available information on CDW data access. A protocol for this review was prepared using the PRISMA-P 2015 Checklist (24) and was preregistered and published within the Open Science Framework (25). The methods of this study are presented in accordance with the Preferred Reporting Items for Systematic Reviews and Meta-Analyses (PRISMA) Statement where applicable (26).

### Search and selection

First, to review the scientific literature, the search engines PubMed, Web of Science, ACM Digital Library, CINAHL and IEEE Xplore were queried systematically. The PRESS checklist served to ensure the inclusion of essential elements in the search strategy (27). The search terms were developed iteratively by piloting combinations of key words and MeSH terms in PubMed and the results were assessed for the inclusion of known representative literature. This process resulted in the combination of key words and MeSH terms of the search strategy presented in table 1. The piloting was performed in October 2018, and the final search was conducted in November 2018.

**Table 1.**
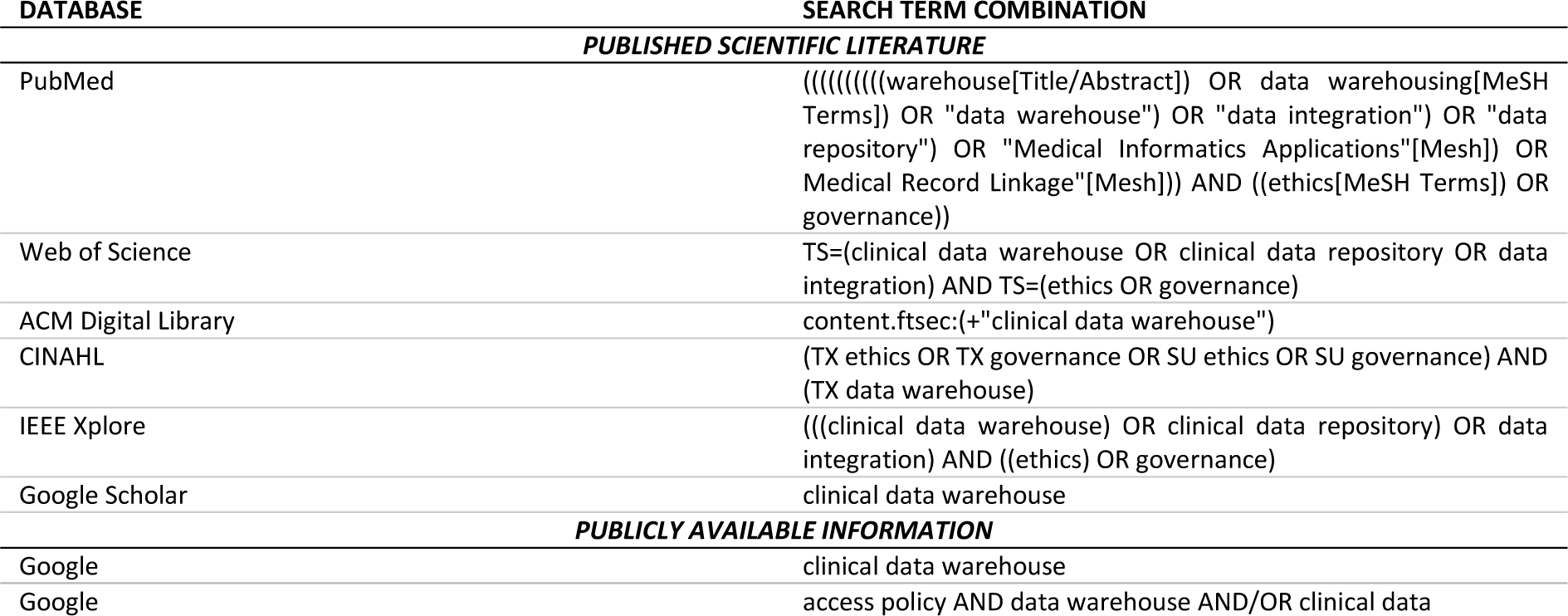
Search strategy.

The retrieved scientific literature was imported into Endnote (Version X9.1) and screened for eligibility using RAYYAN (28). Two authors (EP, HL) independently screened all records and checked for eligibility and inclusion. In order to be included, scientific literature needed to explicitly report the development, implementation or maintenance of a CDW and provide information on the governance of data access and use. Further inclusion criteria were the language of the publication (English, German, or French) and the type of CDW. Only publications that described large-scale CDW (e.g., academic medical centers, larger hospitals) that process data collected from routine health care, and disease-general CDW (not, for example, CDW specific to diabetes) were included. We did not set any restriction based on the date of publication of the source.

After screening the first 100 references, we compared the initial results and discussed the appropriateness of the inclusion and exclusion criteria. In screening the full body of literature, 42 conflicting ratings occurred, all of which were resolved by discussion.

In addition, the search engine Google Scholar was consulted using the key search term elaborated in table 1. The first 200 hits sorted by relevance were included. Finally, in a reference check, the references of all included publications were screened to check for additional literature that had not been captured by our search strategy.

Second, for the publicly available information, a web search was conducted via Google.de to identify the online presence of CDW. The first 100 hits sorted by relevance (by default) were screened for CDW websites (see table 1). There, we searched for publicly available information on data access governance, such as linked access policy documents or access and use criteria addressed directly on the web pages. Further, we searched specifically for the web presences of the CDW of the included literature to check for online available policies.

In addition, Google.de was separately searched for “access policies” (see table 1). All web searches were performed using Google.de in a private browsing mode of the Mozilla Firefox web browser, with all cookies deleted and all private accounts logged out of prior to the search.

### Analysis and synthesis

A qualitative thematic analysis was applied to all included literature (29). Three main thematic categories were developed through an inductive strategy. The main categories were then applied to the literature, and the relevant text passages were coded using MAXQDA 2018 (30). We sensitively extracted all the information potentially relevant to data access and use governance. The findings were then clustered into a matrix containing the spectrum of thematic categories and subcategories derived from the literature. As this review did not focus on the results of interventions, we did not anticipate the need for methods to minimize the risk of bias.

## RESULTS

The systematic scientific literature search retrieved a total of 4249 references. After duplicates were removed, 4133 references were screened. Fifty-one references were then included in the full-text screening, 19 of which fulfilled the criteria and were included in the final analysis. Four additional publications were included via the Google Scholar searches. With the additional web search strategy applied in this study, we found 14 CDW web pages, none of which contained publicly available information on data access governance, such as linked access policy documents or access and use criteria (see supplement table 1 for a list of the CDW web pages). We found one access policy that outlined the governance arrangements of CDWs and fulfilled our inclusion criteria, as determined by checking the web pages of the 23 CDWs of the publications included in the study. The reference check of all included publications did not yield additional literature or documents. (11, 14, 15, 31-50). One intermediate outcome of our study is therefore that only one out of 37 CDW (14 in website search, 27 from scientific literature) had a policy retrievable from their website.

A final set of 24 documents (23 publications and one access policy) was included for the qualitative thematic analysis. See the PRISMA flow diagram in figure 1 for details. All included publications were in English. Twenty-two publications were published in peer-reviewed journals, and one was included in conference proceedings. See supplement table 2 for details on all included documents.

**Figure 1.**
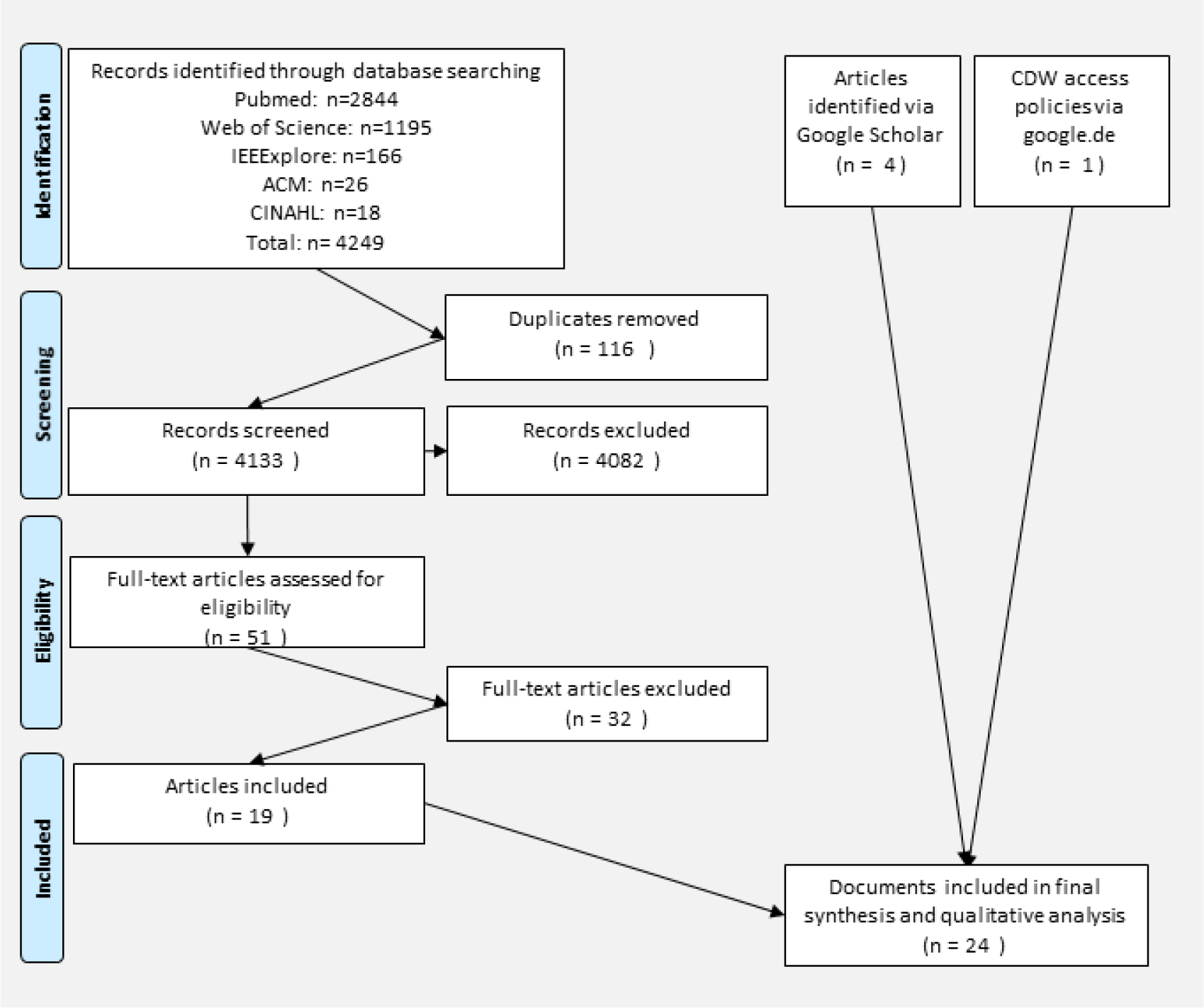
Systematic search flow diagram based on PRISMA.

The primary outcome of this systematic review is a matrix containing the qualitative spectrum of criteria and procedures applied in data access and use governance in CDW (see table 2). The matrix is divided into three main categories: (1) requirements, including the subcategories recipient, reuse and formal requirements, (2) structures and procedures, including the subcategories review bodies and values, and (3) access, including the subcategory access limitations.

**Table 2.**
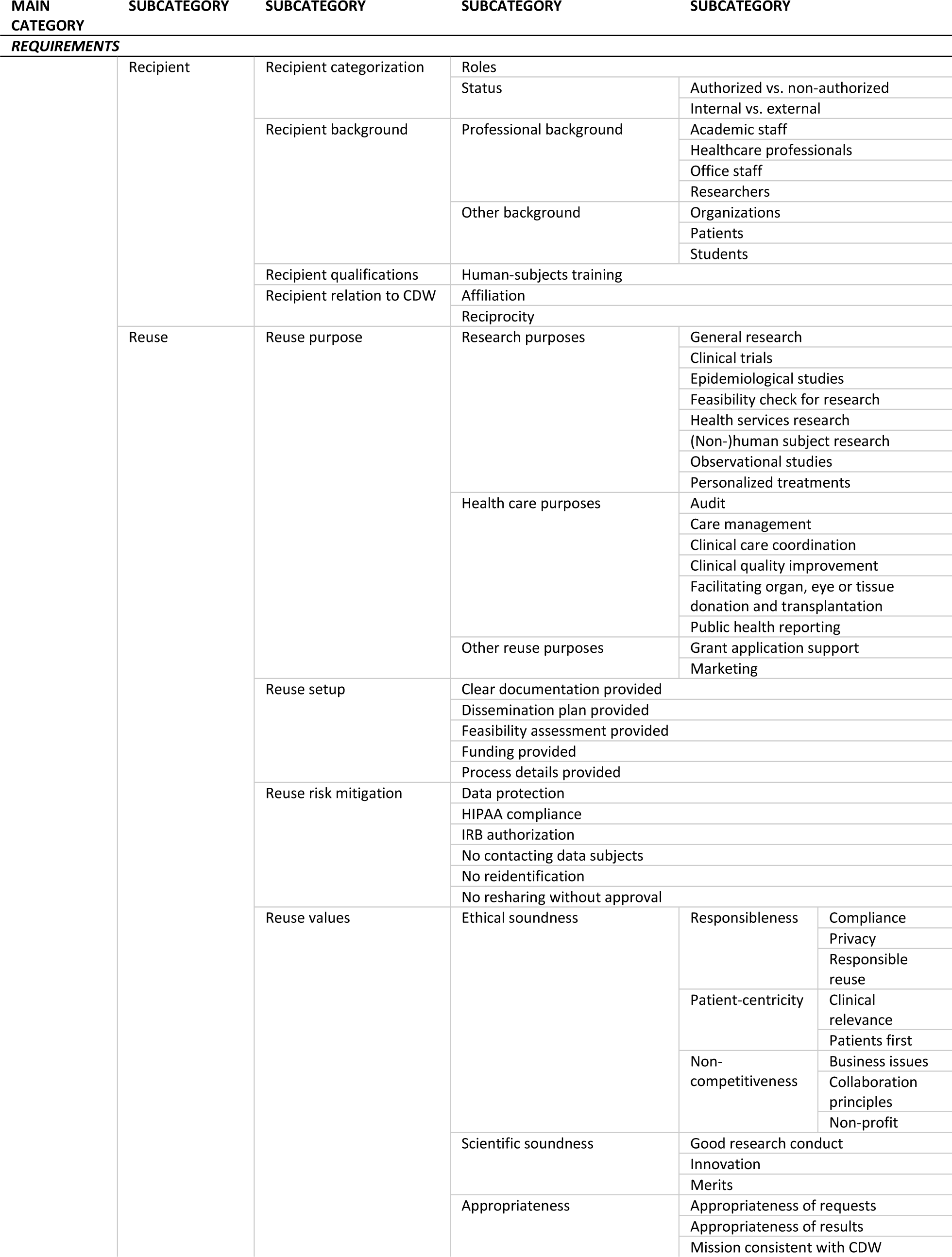
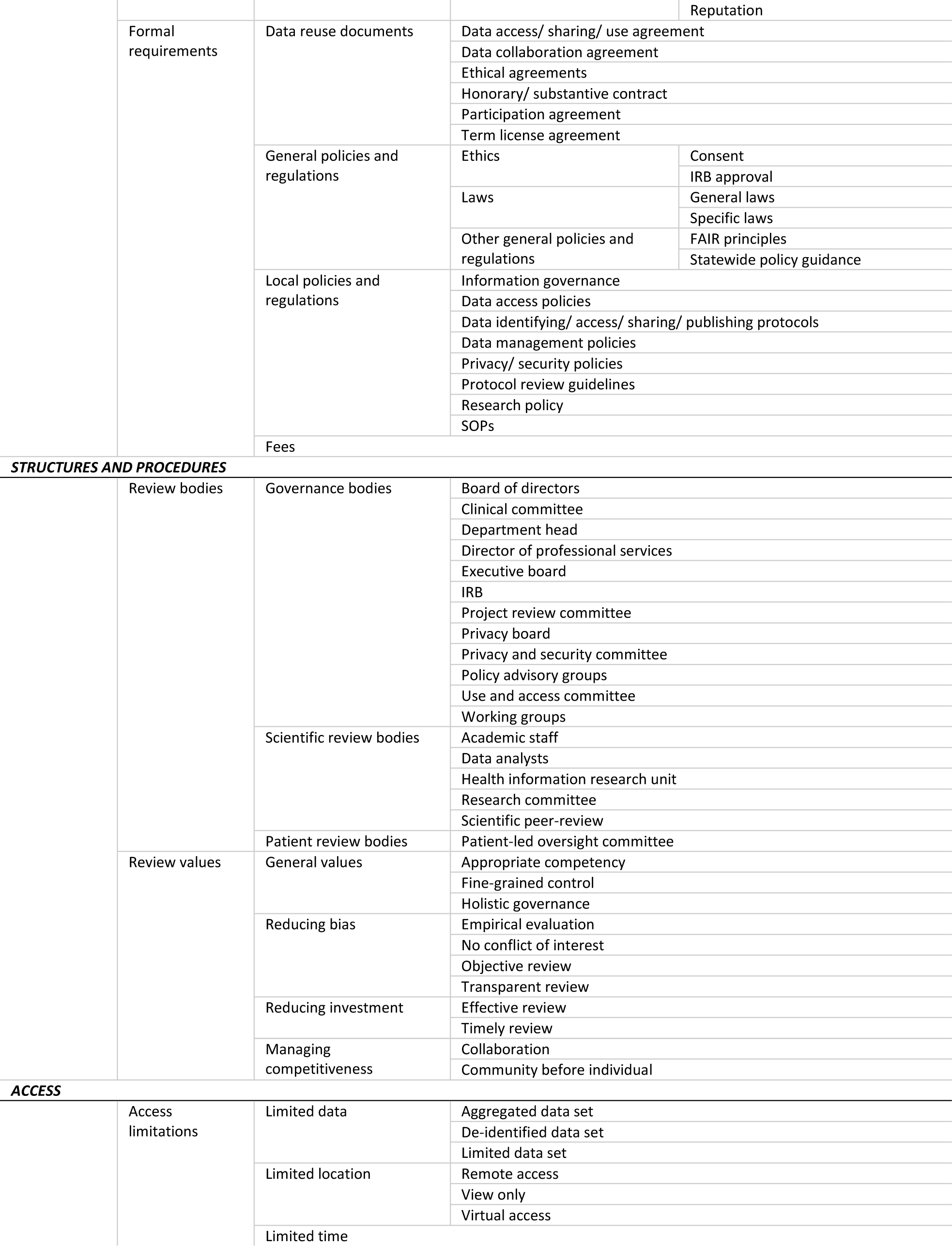
Qualitative spectrum of criteria and procedures in data use and access governance in CDW.

All six subcategories are further split into more detailed subcategories addressing particular aspects relating to data access and use. See table 2 for the full set of categories and subcategories. An expanded table giving exemplary quotes from the included literature for all subcategories is available as an online supplement (supplement table 3).

The policy retrieved contributed substantially to our results (51). What stood out was the clarity of definitions and terms in the document. However, while providing a detailed list for a data reuse “application”, it does not expand on what will ultimately be decisive for a data access and use decision: “(…) compliance with such other criteria as the Research Committee deems appropriate for the Research in question and the protection of Healthix.” (p.23) and “If deemed feasible, the application will then be presented to the Research Committee for review and final action, and such final action shall be communicated in writing to the requesting Researcher.” (p.25) (51). In contrast, the scientific literature was lacking uniformity in terminology and precision overall. It also rarely provided clear indication of whether a certain aspect directly contributes to a data access and use decision.

In the following, we present a brief definition of the themes to facilitate the understanding of the matrix. Figure 2 serves as an overview illustrating the relationship between requirements, structures and procedures, and access.

**Figure 2.**
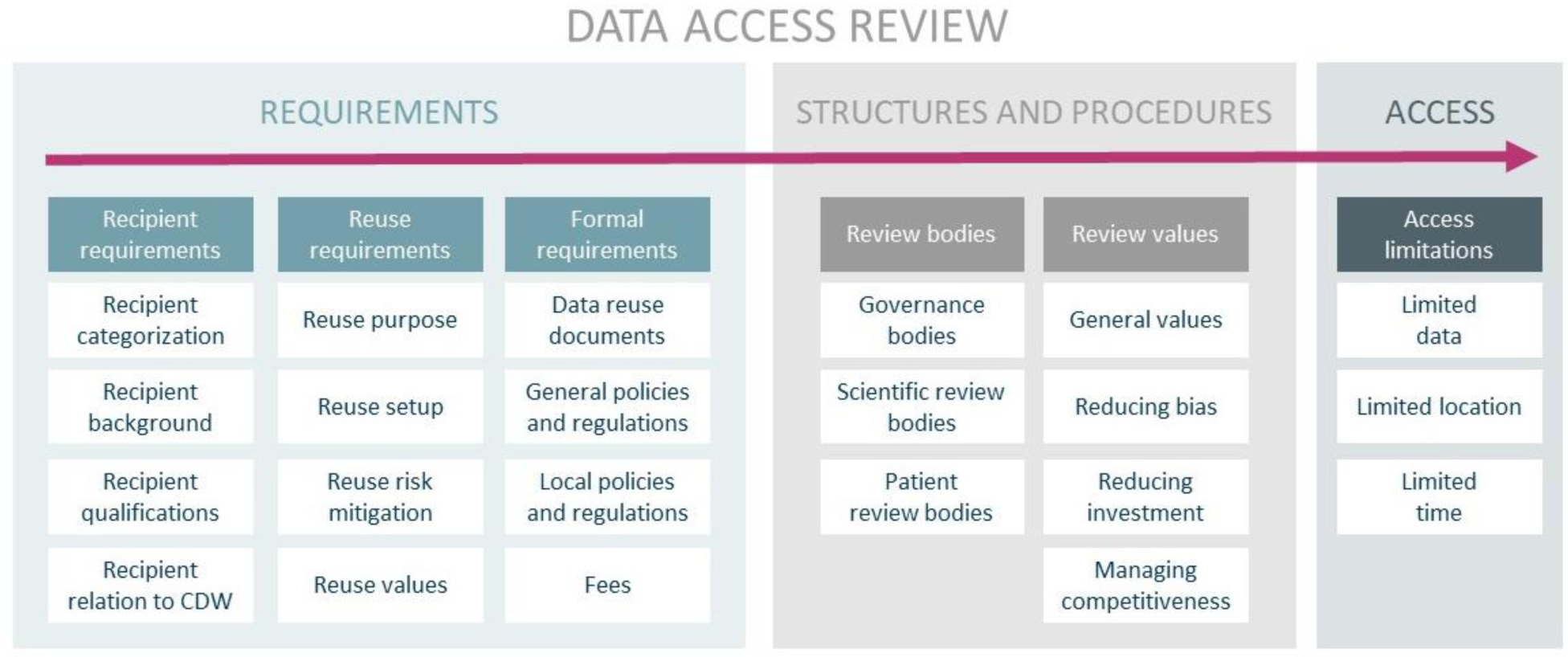
Data access review: Overview of the main categories and two subcategory levels.

### 1. Requirements

Under the main category “requirements”, we subsume all prerequisites linked to the data request. We distinguish between the requirements relating to the potential data recipient (recipient requirements), the requirements relating to the data reuse (reuse requirements) and the requirements relating to documents, policies or other formalities (formal requirements).

### 1.1 Recipient requirements

The recipient requirements include various definitions of potential data recipients, organized by their formal categorization, their background, their qualifications or their relation to the CDW.

#### Recipient categorization

Recipients can be categorized in different manners. On a general level, these categories can be divided into roles and status. Role represents a specific combination of attributes defined in detail by the respective CDW (e.g. background and qualifications combined). Status describes the presence or lack of one concrete attribute (e. g. authorized or not, internal or not).

#### Recipient background

On a more detailed level, potential data recipients are characterized by their background. This can be a specific professional background or another background relevant to the context of the CDW.

#### Recipient qualifications

Potential data recipients can be required to have certain qualifications in order to access the data, meaning that they have skills that enable them to use the data as required by the CDW.

#### Recipient relation to CDW

The potential data recipient’s relationship to the CDW might play a role in the data request. Some CDW might only agree to data access when a prior relation to the CDW or to the data provided to the CDW has been established.

### 1.2 Reuse requirements

The reuse requirements reflect different aspects of the planned data reuse that might influence access and use decisions.

#### Reuse purposes

Reuses can be described by their intended purpose, meaning the proposed objective of the data reuse. We intentionally excluded primary uses such as treatment and the closely related purposes of payment and insurance coverage review as presented in the policy document (51), as this review focuses on data reuse.

#### Reuse setup

Reuse setup describes organizational and practical level preparations for the data reuse that need to be provided for a data request.

#### Reuse risk mitigation

Reuse risk mitigation describes the potentially required commitments of the data reuser that serve to minimize the risk to the data subject stemming from the reuse of data.

#### Reuse values

Reuse values are requirements to the quality and integrity of the reuse request concerning ethical or scientific soundness, or appropriateness. *Ethical soundness* can be further subdivided into responsibleness, patient-centricity and non-competitiveness.

### 1.3 Formal requirements

Formal requirements are documents, policies and regulations which potential data recipients might need to sign, respect or agree to for a positive data access and use decision.

#### Data reuse documents

Data reuse documents describe mutual agreements and contracts specific to the reuse of the data.

#### General policies and regulations

This section describes fundamental laws and policies whose requirements are not specific to the CDW but more general.

#### Local policies and regulations

In contrast to the general policies and regulations, this subcategory comprises the policies and regulations where the content would be expected to vary strongly depending on the local particularities.

#### Fees

The data access and use decisions might also rely on the potential recipient’s agreement to paying a fee.

### 2. Structure and procedures

Under the main category “structure and procedures”, the governance structures and procedures are subsumed which need to be in place to provide for a review of the data request and the subsequent data access and use decision. They are divided into review bodies who are exercising the review, and review values underlying review procedures and decisions.

### 2.1 Review bodies

Review bodies reflect the implementation of designated governance bodies, which is commonplace in all assessed CDW. Here, we do not distinguish between boards, groups and individual positions, as the hierarchies and directions are not always transparent.

#### Governance bodies

This section comprises a variety of rather general governance bodies involved in the data reuse review.

#### Scientific review bodies

As a more specialized review body type, scientific review bodies describe those dealing with research questions.

#### Patient review bodies

Another type of specific review bodies are those where patients are involved in data access and use decisions.

### 2.2 Review values

Review bodies might follow certain review values when deciding data access and use requests which are subsumed in the following.

#### General values

General values describe values driving the CDW’s overall data access governance structure and procedures.

#### Reducing bias

This section encompasses the measures taken to reduce bias in the data request review.

#### Reducing investment

Reducing investments reflects aspects that save valuable resources, such as time and effort.

#### Managing competitiveness

Managing competitiveness describes an approach that places common interest over individual interest.

## 3. Access

Under the main category “access”, we subsume data access which can be granted with varying limitations based on the previously described requirements or particularities of the CDW.

### 3.1 Access limitations

Access limitations can take on different forms and restrict either data itself, location or the access time.

#### Limited data

Data access can also be limited in itself, if only a specific part of the requested data is provided, or if it is only provided in a specific form that might be reduced in information.

#### Limited location

Another possible limitation in access can be the location, where data can only be accessed from specific places.

#### Limited time

Some CDW might restrict the access to data to a certain timeframe or number of accesses.

## DISCUSSION

This systematic review aimed to assess the spectrum of criteria and procedures that surround data access and use decisions in CDW. A core finding is the lack of concrete information available in the scientific literature and the lacking public accessibility of CDW data access policies.

### Focus of the current scientific discussion

While there is a solid corpus of scientific literature on the implementation and maintenance of CDW, the governance of data access is only marginally addressed. In many cases, the publications retrieved mention the relevance of data access and use without clarifying the details on the handling in practice. Many of the publications instead focus on technical aspects of data governance. Since the practical aspects of data access and use only become relevant when a significant amount of data is available for sharing, it is reasonable that CDW might approach technical issues first, which also require considerable effort and resources.

However, international recommendations on the ethics of health data sharing also emphasize the importance of procedural (e.g. independent review) and substantial aspects (e.g. criteria for data access decisions) of data access governance (20, 52). The design and implementation of such sound governance arrangements for data access and use are nonetheless challenging (53).

Procedural aspects might be comparatively straightforward. Despite the heterogeneity of the denominations and of the composition of governance bodies, based on our study such procedural aspects seem to be addressed appropriately in the scientific literature.

Substantial aspects can be considered more demanding. The elaboration of concise access criteria satisfying all stakeholders and enabling all foreseen data reuses is a highly complex task. Considering the amount of public funding invested in the establishment of CDW, sharing details on the conceptualization and the access and use criteria themselves with the scientific community and the public is all the more important. This could significantly improve the efficiency of funding devoted to CDW.

### Accessibility of policies, and access and use criteria

In publications on transparent access and use governance, we would expect to find concrete and decisive criteria on the basis of which data access requests are weighted and judged. While the existence of more concrete policies is mentioned in the publications included, they could not be retrieved in our separate search except for one.

In the additional web search for publicly available access and use policy documents, our strategy was reduced to simpler search terms (see table 1). A more refined search string could have potentially produced more policy documents. While this can be considered a limitation of our study, it also emulates how researchers and the public would most likely search for information on CDW.

To our knowledge, other than for biobanks (54) there are no public registries for CDW that would help access such information. It therefore appears that the access to those policies is limited, both by the CDW themselves and through a lack of appropriate search instruments. This is an important finding, as it reveals significant shortcomings in the compliance of CDW with the internationally agreed upon and accepted ethical standards of transparency, as required, for instance, by the WMA (20).

### Transparency of criteria and its ethical value

Through our screening, we were able to extract potential criteria for decisions on data access and use. However, most of these criteria were not explicitly described as being decisive for a data request. Criteria applicable to the handling of data access requests are often provided in a rather unspecific and unstructured manner. They are not addressed in a separate designated section of the publications but are mentioned secondarily or need to be deduced from other information. Even the more explicit access and use criteria offer considerable leeway for interpretation and are therefore difficult to translate into practice. In addition, the denomination of requirements, structures and procedures varies highly between CDW and often lacks a more distinct definition. This can be detrimental to the harmonization and effectiveness of the CDW’s objectives.

The need for more clarity and standards on conditions for access and use must also be considered an ethical issue. While information transparency is not an ethical principle on its own, it can certainly become ethically relevant, for example when ethical principles such as informed consent or accountability depend on that information (55).

One could argue that individuals consenting to their data being stored and processed by a CDW rely on the CDW’s governance structures to execute the data reuse, first, according to their given consent, and second, with their best interests in mind beyond the consent. In their publication on the governance of biobanks, the German Ethics Council states that in order to compensate for the lack of precision at the time of consenting, donors should have the capacity to trace the governance of sample and data transfer. The specific purposes of reuse should therefore be publicly accessible in “a clear, generally intelligible and up-to-date account” (p.44) (56). This should be considered analogous for CDW. Transparency can therefore be considered a means to involve the data subjects and to give them a higher degree of data control. While some CDW embrace the direct involvement of patients in governance processes, for example in patient-led oversight committees, such measures are likely to improve transparency on a more individual level.

Particularly important, moreover, is the role of transparency as a requirement for setting up accountability mechanisms. Only clear conditions and attributions allow for the evaluation and execution of compliance with ethical and other norms. Ultimately, accountability also serves to increase public trust in data reuse (19).

Increased trust in CDW is also likely to have a positive effect on the future willingness to share data (57). In contrast, the limited availability of information in the scientific literature and on CDW websites might prove to be detrimental. On a larger scale, this could weaken the potential of LHCS and other promising outcomes of secondary uses of data, such as precision medicine or comparative effectiveness studies, as they rely on the public’s investment in research.

### Return on public investment

Expectations placed on data-driven approaches in biomedical research and health care are high. Accordingly, increased efforts are being made internationally to develop and implement IT infrastructures that facilitate high-quality data access and use. In Germany, for instance, the Medical Informatics Initiative funding scheme was launched by the Federal Ministry of Research and Education (BMBF) in 2016 and aims to foster collaborative data use by investing 150 million Euros into the development of robust IT solutions (58).

This considerable allocation of funding, however, is generally based on the assumption of a return on public investment. Normative concepts and recommendations for the ethical conduct of data-driven approaches in medical research and health care have been developed and almost unanimously highlight the importance of data access (20, 59-61). However, with an increasing amount of data requests for data-driven approaches, the case-by-case decisions as they appear to be reflected by our study will need to be replaced by a more automatized approach. In order to implement such automatization, the criteria and their weighting must be unambiguously clear.

Both, promoting scientific research and improving health care delivery, are very much in the public interest. While the public is supportive of the reuse of data for research purposes [35], it is the transparency of data access and use governance that might ensure their long-term support and trust through efficient reuse, informed consent, and accountability.

## CONCLUSION

The results of this review may contribute to the development of practice-oriented minimal standards for the governance of data access and use, which could also contribute to more harmonization, efficiency, and effectiveness.

The study can further serve as an indicator for the practical implementation of (soft) regulations and statements of intent for data access. It may inform researchers and physicians, funders, and hospital administration staff involved in the design and implementation of CDW policies on data access and use. In the long run, more emphasis should be placed on the practice evaluation of CDW governance to assess compliance with ethical standards and to identify practical issues (62). The latter, in turn, will be relevant for further policy development.

As digitalization and data-driven approaches in health research and health care are rapidly evolving, the current governance practices will require broader implementation, evaluation, and improvement to keep up with ongoing developments and challenges.

## Data Availability

An extensive part of the data analyzed during this study is included in this published article and its supplementary files (Supplement Table 3). The raw dataset analyzed during the current study is not publicly available in full due to copyright restrictions but is available from the corresponding author on reasonable request.

## LIST OF ABREVIATIONS

BMBF: German Federal Ministry of Education and Research
CDW: Clinical data warehouse
EHR: Electronic health records
LHCS: Learning health care systems
PRISMA: Preferred Reporting Items for Systematic Reviews and Meta-Analyses
WMA: World Medical Association

## DECLARATIONS

### Ethics approval and consent to participate

Not applicable.

### Consent for publication

Not applicable.

### Authors’ contributions

DS and HL conceived the work. HL and EP contributed to the design, the acquisition, the analysis, and the interpretation of the data, as well as the draft and revision of the work. All three authors consented on the final version of the paper.

## Acknowledgments

We thank Delwen Franzen for her helpful comments on our draft.

## REFERENCES

1. Adler-Milstein J, DesRoches CM, Kralovec P, Foster G, Worzala C, Charles D, et al. Electronic health record adoption in US hospitals: progress continues, but challenges persist. Health affairs. 2015;34(12):2174–80.

2. Longhurst CA, Harrington RA, Shah NH. A ‘green button’ for using aggregate patient data at the point of care. Health affairs (Project Hope). 2014;33(7):1229–35.

3. Meystre SM, Lovis C, Burkle T, Tognola G, Budrionis A, Lehmann CU. Clinical Data Reuse or Secondary Use: Current Status and Potential Future Progress. Yearbook of medical informatics. 2017;26(1):38–52.

4. Diamond CC, Mostashari F, Shirky C. Collecting and sharing data for population health: a new paradigm. Health affairs (Project Hope). 2009;28(2):454–66.

5. Friedman CP, Wong AK, Blumenthal D. Achieving a nationwide learning health system. Science translational medicine. 2010;2(57):57cm29.

6. Jensen PB, Jensen LJ, Brunak S. Mining electronic health records: towards better research applications and clinical care. Nature Reviews Genetics. 2012;13:395.

7. Emanuel EJ, Wachter RM. Artificial Intelligence in Health Care: Will the Value Match the Hype? Jama. 2019.

8. Khoumbati K, Themistocleous M. Integrating the IT infrastructures in healthcare organisations: a proposition of influential factors. The Electronic Journal of e-Government. 2006;4(1):27–36.

9. Dixon BE, Vreeman DJ, Grannis SJ. The long road to semantic interoperability in support of public health: experiences from two states. J Biomed Inform. 2014;49:3–8.

10. Schneeweiss S. Learning from Big Health Care Data. 2014;370(23):2161–3.

11. Chute CG, Beck SA, Fisk TB, Mohr DN. The Enterprise Data Trust at Mayo Clinic: a semantically integrated warehouse of biomedical data. J Am Med Inform Assoc. 2010;17(2):131–5.

12. Shin S-Y, Kim WS, Lee J-H. Characteristics desired in clinical data warehouse for biomedical research. Healthcare informatics research. 2014;20(2):109–16.

13. Evans RS, Lloyd JF, Pierce LA. Clinical use of an enterprise data warehouse. AMIA Annual Symposium proceedings AMIA Symposium. 2012;2012:189–98.

14. Foran DJ, Chen W, Chu H, Sadimin E, Loh D, Riedlinger G, et al. Roadmap to a Comprehensive Clinical Data Warehouse for Precision Medicine Applications in Oncology. Cancer informatics. 2017;16:1176935117694349.

15. Horvath MM, Winfield S, Evans S, Slopek S, Shang H, Ferranti J. The DEDUCE Guided Query tool: Providing simplified access to clinical data for research and quality improvement. Journal of Biomedical Informatics. 2011;44(2):266–76.

16. Rosenbaum S. Data governance and stewardship: designing data stewardship entities and advancing data access. Health services research. 2010;45(5 Pt 2):1442–55.

17. Ford E, Boyd A, Bowles JK, Havard A, Aldridge RW, Curcin V, et al. Our data, our society, our health: A vision for inclusive and transparent health data science in the United Kingdom and beyond. Learning Health Systems. 2019:e10191.

18. Vayena E, Dzenowagis J, Brownstein JS, Sheikh A. Policy implications of big data in the health sector. Bulletin of the World Health Organization. 2018;96(1):66.

19. Blasimme A, Fadda M, Schneider M, Vayena E. Data Sharing For Precision Medicine: Policy Lessons And Future Directions. Health affairs (Project Hope). 2018;37(5):702–9.

20. aWorld Medical Association. WMA declaration of Taipei on ethical considerations regarding health databases and biobanks. WMA; 2016.

21. Langhof H, Kahrass H, Sievers S, Strech D. Access policies in biobank research: what criteria do they include and how publicly available are they? A cross-sectional study. European journal of human genetics : EJHG. 2017;25(3):293–300.

22. Langhof H, Kahrass H, Illig T, Jahns R, Strech D. Current practices for access, compensation, and prioritization in biobanks. Results from an interview study. European journal of human genetics : EJHG. 2018;26(11):1572–81.

23. Holmes JH, Elliott TE, Brown JS, Raebel MA, Davidson A, Nelson AF, et al. Clinical research data warehouse governance for distributed research networks in the USA: a systematic review of the literature. Journal of the American Medical Informatics Association : JAMIA. 2014;21(4):730–6.

24. Moher D, Shamseer L, Clarke M, Ghersi D, Liberati A, Petticrew M, et al. Preferred reporting items for systematic review and meta-analysis protocols (PRISMA-P) 2015 statement. 2015;4(1):1.

25. Governance of Data Sharing. [Internet]. Open Science Framework 2018. Available from: https://osf.io/6w4n5/.

26. Moher D, Liberati A, Tetzlaff J, Altman DGJAoim. Preferred reporting items for systematic reviews and meta-analyses: the PRISMA statement. 2009;151(4):264–9.

27. McGowan J, Sampson M, Salzwedel DM, Cogo E, Foerster V, Lefebvre C. PRESS Peer Review of Electronic Search Strategies: 2015 Guideline Statement. Journal of clinical epidemiology. 2016;75:40–6.

28. Ouzzani M, Hammady H, Fedorowicz Z, Elmagarmid A. Rayyan-a web and mobile app for systematic reviews. Systematic reviews. 2016;5(1):210.

29. Braun V, Clarke VJQrip. Using thematic analysis in psychology. 2006;3(2):77–101.

30. VERBI Software GmbH. MAXQDA, Software for qualitative data anaylsis Berlin, Germany: VERBI Software Consult Sozialforschung GmbH; [Available from: http://www.maxqda.com/products/maxqda.

31. Bouzillé G, Westerlynck R, Defossez G, Bouslimi D, Bayat S, Riou C, et al. Sharing health big data for research-A design by use cases: the INSHARE platform approach. Stud Health Technol Inform. 2017(245):303–7.

32. Des Jardins TR. The keys to governance and stakeholder engagement: the southeast michigan beacon community case study. EGEMS (Washington, DC). 2014;2(3):1068.

33. Fleischman W, Lowry T, Shapiro J. The visit-data warehouse: enabling novel secondary use of health information exchange data. EGEMS (Washington, DC). 2014;2(1):1099.

34. Ford DV, Jones KH, Verplancke JP, Lyons RA, John G, Brown G, et al. The SAIL Databank: building a national architecture for e-health research and evaluation. BMC health services research. 2009;9:157.

35. Grant A, Moshyk A, Diab H, Caron P, de Lorenzi F, Bisson G, et al. Integrating feedback from a clinical data warehouse into practice organisation. International journal of medical informatics. 2006;75(3-4):232–9.

36. Haarbrandt B, Schreiweis B, Rey S, Sax U, Scheithauer S, Rienhoff O, et al. HiGHmed - An Open Platform Approach to Enhance Care and Research across Institutional Boundaries. Methods Inf Med. 2018;57(S 01):e66–e81.

37. Hazlehurst BL, Kurtz SE, Masica A, Stevens VJ, McBurnie MA, Puro JE, et al. CER Hub: An informatics platform for conducting comparative effectiveness research using multi-institutional, heterogeneous, electronic clinical data. International journal of medical informatics. 2015;84(10):763–73.

38. Jannot AS, Zapletal E, Avillach P, Mamzer MF, Burgun A, Degoulet P. The Georges Pompidou University Hospital Clinical Data Warehouse: A 8-years follow-up experience. International journal of medical informatics. 2017;102:21–8.

39. Laws R, Gillespie S, Puro J, Van Rompaey S, Quach T, Carroll J, et al. The Community Health Applied Research Network (CHARN) Data Warehouse: a Resource for Patient-Centered Outcomes Research and Quality Improvement in Underserved, Safety Net Populations. EGEMS (Washington, DC). 2014;2(3):1097.

40. Lowe HJ, Ferris TA, Hernandez PM, Weber SC. STRIDE--An integrated standards-based translational research informatics platform. AMIA Annu Symp Proc. 2009;2009:391–5.

41. Liu J, Erdal S, Silvey SA, Ding J, Riedel JD, Marsh CB, et al. Toward a fully de-identified biomedical information warehouse. AMIA Annu Symp Proc. 2009;2009:370–4.

42. Perera G, Broadbent M, Callard F, Chang CK, Downs J, Dutta R, et al. Cohort profile of the South London and Maudsley NHS Foundation Trust Biomedical Research Centre (SLaM BRC) Case Register: current status and recent enhancement of an Electronic Mental Health Record-derived data resource. BMJ open. 2016;6(3):e008721.

43. Prasser F, Kohlbacher O, Mansmann U, Bauer B, Kuhn KA. Data Integration for Future Medicine (DIFUTURE). Methods Inf Med. 2018;57(S 01):e57–e65.

44. Prokosch HU, Acker T, Bernarding J, Binder H, Boeker M, Boerries M, et al. MIRACUM: Medical Informatics in Research and Care in University Medicine. Methods Inf Med. 2018;57(S 01):e82-e91.

45. Ross TR, Ng D, Brown JS, Pardee R, Hornbrook MC, Hart G, et al. The HMO Research Network Virtual Data Warehouse: A Public Data Model to Support Collaboration. EGEMS (Washington, DC). 2014;2(1):1049.

46. Stark PC, Kalenderian E, White JM, Walji MF, Stewart DC, Kimmes N, et al. Consortium for oral health-related informatics: improving dental research, education, and treatment. Journal of dental education. 2010;74(10):1051–65.

47. Turley CB, Obeid J, Larsen R, Fryar KM, Lenert L, Bjorn A, et al. Leveraging a Statewide Clinical Data Warehouse to Expand Boundaries of the Learning Health System. EGEMS (Washington, DC). 2016;4(1):1245.

48. Van Eaton EG, Devlin AB, Devine EB, Flum DR, Tarczy-Hornoch P. Achieving and sustaining automated health data linkages for learning systems: barriers and solutions. EGEMS (Washington, DC). 2014;2(2):1069.

49. Walji MF, Kalenderian E, Stark PC, White JM, Kookal KK, Phan D, et al. BigMouth: a multi-institutional dental data repository. J Am Med Inform Assoc. 2014;21(6):1136–40.

50. Winter A, Staubert S, Ammon D, Aiche S, Beyan O, Bischoff V, et al. Smart Medical Information Technology for Healthcare (SMITH). Methods Inf Med. 2018;57(S 01):e92–e105.

51. Healthix. Heathix: Security Policies and Procedures 2019 [Available from: https://healthix.org/who-we-are/healthix-policies/.

52. Council of Europe. Recommendation CM/Rec(2016)6 of the Committee of Ministers to member States on research on biological materials of human origin 2016 [Available from: https://search.coe.int/cm/Pages/result_details.aspx?ObjectId=090000168064e8ff.

53. Ohno-Machado L. To share or not to share: that is not the question. Science translational medicine. 2012;4(165):165cm15–cm15.

54. Holub P, Swertz M, Reihs R, van Enckevort D, Müller H, Litton J-E. BBMRI-ERIC directory: 515 biobanks with over 60 million biological samples. Biopreservation and biobanking. 2016;14(6):559–62.

55. Turilli M, Floridi L. The ethics of information transparency. Ethics and Information Technology. 2009;11(2):105–12.

56. Ethikrat N. Human biobanks for research. Berlin: The German National Ethics Council. 2010.

57. Vayena E, Blasimme A. Biomedical Big Data: New Models of Control Over Access, Use and Governance. Journal of bioethical inquiry. 2017;14(4):501–13.

58. Gehring S, Eulenfeld R. German Medical Informatics Initiative: Unlocking data for research and health care. Methods of information in medicine. 2018;57(S 01):e46–e9.

59. OECD. Recommendation of the OECD Council on Health Data Governance 2017 [Available from: https://www.oecd.org/health/health-systems/Recommendation-of-OECD-Council-on-Health-Data-Governance-Booklet.pdf.

60. Nuffield Council on Bioethics. The Collection, Linking and Use of Data in Biomedical Research and Health Care: Ethical Issues: Nuffield Council on Bioethics; 2015.

61. Deutscher Ethikrat. Big Data und Gesundheit–Datensouveränität als informationelle Freiheitsgestaltung. 2017.

62. Langhof H, Schwietering J, Strech D. Practice evaluation of biobank ethics and governance: current needs and future perspectives. Journal of medical genetics. 2018.

